# Loss of Taste and Smell as Distinguishing Symptoms of COVID-19

**DOI:** 10.1101/2020.05.13.20101006

**Authors:** Patrick Dawson, Elizabeth M. Rabold, Rebecca L. Laws, Erin E. Conners, Radhika Gharpure, Sherry Yin, Sean A. Buono, Trivikram Dasu, Sanjib Bhattacharyya, Ryan P. Westergaard, Ian W. Pray, Dongni Ye, Scott A. Nabity, Jacqueline E. Tate, Hannah L. Kirking

## Abstract

Olfactory and taste dysfunctions have emerged as symptoms of COVID-19. Among individuals with COVID-19 enrolled in a household study, loss of taste and/or smell was the fourth most commonly reported symptom (26/42; 62%), and among household contacts, it had the highest positive predictive value (83%; 95% CI: 55–95%) for COVID-19. These findings support consideration of loss of taste and/or smell in possible case identification and testing prioritization for COVID-19.

## Main

The global coronavirus disease 2019 (COVID-19) pandemic caused by severe acute respiratory syndrome coronavirus 2 (SARS-CoV-2) has prompted robust public health investigations as healthcare workers and epidemiologists attempt to characterize the disease course. Early published reports on symptoms associated with infection focused primarily on cases with hospitalizations and emergency department visits. Among these patients with moderate to severe disease, fever, cough, and shortness of breath were the most frequently described symptoms.^1,2^

Because recent reports have identified additional symptoms associated with SARS-CoV-2 infection, the U.S. Centers for Disease Control and Prevention (CDC) and the Council for State and Territorial Epidemiologists (CSTE) updated their list of symptoms compatible with SARSCoV-2 infection in March 2020.^3,4^ In addition to the classic COVID-19 symptoms of fever, cough, and shortness of breath, loss of taste and smell were added as possible symptoms. Although several reports have described these newly recognized symptoms of loss of taste and smell among individuals diagnosed with COVID-19,^5,6^ few studies have prospectively evaluated these symptoms among close contacts of COVID-19 cases prior to diagnosis and outside of a clinical setting.

In March and April 2020, CDC, in partnership with Wisconsin Department of Health Services and local health departments in Milwaukee County, conducted a household study that included known COVID-19 index cases and their household members. This investigation provided an opportunity to identify household COVID-19 cases and describe their symptom profiles, including loss of taste and smell. This investigation ascertained symptoms prior to diagnosis for household contacts of COVID-19 index cases, similar to how patients might present in a clinical setting early in the course of illness.

We enrolled 90 participants from 26 households, including 26 index cases and 64 household members. Overall, 48 (53%) study participants were male; 69 (77%) were adults ≥18 years old; 41 (46%) were black non-Hispanic/Latino, 37 (41%) were white non-Hispanic/Latino, and 12 (13%) were other races/ethnicities. Preexisting medical conditions were reported by 39 (43%) participants, including asthma or reactive airway disease (n = 13), hypertension (n = 10), and diabetes (n = 8). Upon enrollment, 16/64 (25%) household contacts tested positive for SARSCoV-2, for a total of 42/90 (47%) COVID-19 cases. Median time from symptom onset to enrollment for index cases and symptomatic household cases was 14.5 days [interquartile range (IQR): 10.0–19.0] and 7.0 days (IQR 5.0–7.5), respectively.

Among the 42 individuals with laboratory-confirmed SARS-CoV-2, all (100%) reported at least one symptom (**Table 1**); 38 (90%) reported at least one classic COVID-19 symptom of fever, cough, or shortness of breath. The most frequently reported symptoms were cough (81%), headache (76%), fever (subjective or measured ≥ 100.4°F) (64%), loss of taste and/or smell (62%), nasal congestion (62%), myalgia (57%), and chills (55%).

**Table 1.**
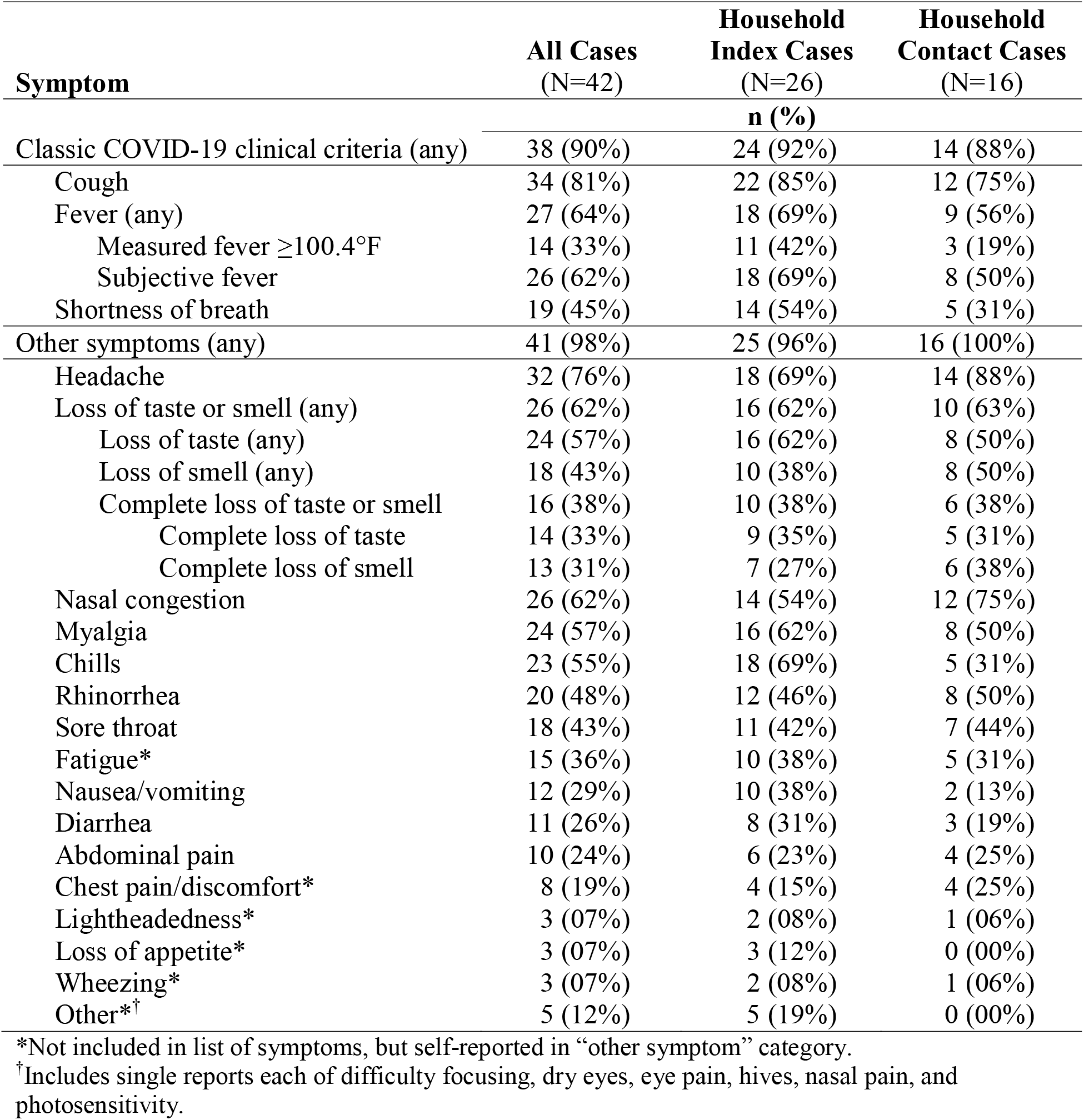
Prevalence of self-reported symptoms among individuals with laboratory-confirmed COVID-19, Milwaukee County, Wisconsin, March–April 2020.

Of the 26 participants with COVID-19 who reported loss of taste and/or smell, loss of taste was reported by 24 participants; 14/24 (58%) describing it as a complete loss. Loss of smell was reported by 18 participants; 13/18 (72%) describing it as a complete loss. There were no significant differences in reporting loss of taste and/or smell by sex, age <18 years versus 18 years or older, race/ethnicity, presence of preexisting medical conditions, or between index cases and household member cases (all p>0.05). Of the 26 participants reporting any loss of taste and/or smell, 9 (35%) reported it in the absence of nasal congestion, including 4 experiencing a complete loss of both taste and smell. Participants with COVID-19 reporting loss of taste and/or smell were more likely to report headache (88% vs. 56%; p = 0.03) but were no more or less likely to report any other symptoms (p>0.05). No participant reported loss of taste and/or smell as the only symptom. Fifty-seven percent experienced loss of taste and/or smell along with at least one classic COVID-19 symptom, 55% with at least one other upper respiratory symptom (nasal congestion, sore throat, and rhinorrhea), and 62% with any other symptom. When loss of taste and/or smell was added to the classic symptoms, 95% of participants with COVID-19 reported at least one of loss of taste and/or smell, fever, cough, and shortness of breath.

Among the 64 household members of COVID-19 index cases, loss of taste and/or smell was reported by 12 individuals, of whom 10 were positive for SARS-CoV-2.

The positive predictive value (PPV) of any loss of taste and/or smell for COVID-19 (83%, 95% CI: 55–95%) was higher than for fever (subjective or measured) and cough, two of the three classic symptoms, and equal to the third, shortness of breath (83%, 95% CI: 44–97%) (**Table 2)**. The PPV for complete loss of taste and/or smell (86%, 95% CI: 49–97%) was the highest among any of the symptoms.

**Table 2.** Positive predictive value of symptoms prior to enrollment testing among household contacts of COVID-19 index cases, Milwaukee County, Wisconsin, March–April 2020.

| **Symptom** | **SARS-CoV-2**<br>**positive**<br>**(N=16)** | **SARS-CoV-2**<br>**negative**<br>**(N=48)** | **Symptom Positive**<br>**Predictive Value**<br>**% (95% CI)** |
| --- | --- | --- | --- |
| Classic COVID-19 clinical criteria (any) | 14 | 14 | 50% (33–67%) |
| Shortness of breath | 5 | 1 | 83% (44–97%) |
| Fever (any) | 9 | 3 | 75% (47–91%) |
| Measured fever $\geq 100.4^{\circ}\text{F}$ | 3 | 1 | 75% (30–95%) |
| Subjective fever | 8 | 2 | 80% (49–94%) |
| Cough | 12 | 13 | 48% (30–67%) |
| Other symptoms (any) | 16 | 29 | 36% (23–50%) |
| Loss of taste or smell (any) | 10 | 2 | 83% (55–95%) |
| Loss of taste (any) | 8 | 2 | 80% (49–94%) |
| Loss of smell (any) | 8 | 1 | 89% (57–98%) |
| Complete loss of taste or smell | 6 | 1 | 86% (49–97%) |
| Complete loss of taste | 5 | 1 | 83% (44–97%) |
| Complete loss of smell | 6 | 1 | 86% (49–97%) |
| Myalgia | 8 | 3 | 73% (43–90%) |
| Nausea/vomiting | 2 | 1 | 67% (21–94%) |
| Chills | 5 | 3 | 63% (31–86%) |
| Nasal congestion | 12 | 9 | 57% (37–76%) |
| Abdominal pain | 4 | 4 | 50% (22–78%) |
| Headache | 14 | 16 | 47% (30–64%) |
| Sore throat | 7 | 10 | 41% (22–64%) |
| Rhinorrhea | 8 | 13 | 38% (21–59%) |
| Diarrhea | 3 | 10 | 23% (08–50%) |

In this household-based population of individuals with COVID-19, which included mildly symptomatic individuals who otherwise may not have been tested according to contemporaneous public health guidance,^7^ loss of taste and/or smell was reported by more than three of every five individuals with confirmed COVID-19. Among the population of 64 household contacts of COVID-19 index cases, it had the highest positive predictive value for COVID-19 of all symptoms, only matched by shortness of breath. Prevalence of SARS-CoV-2 infection among household members of index cases was high, at 25%; therefore, PPV estimates may not be generalizable in populations with different background prevalence. However, our findings may be particularly relevant for screening individuals in close contact with known cases. When compared to most other symptoms, loss of taste and/or smell appeared highly predictive of SARS-CoV-2 infection and was more predictive than cough.

Nasal congestion alone is unlikely to explain the taste and smell alterations, as one-third of patients reporting loss of taste and/or smell did not report nasal congestion; other analyses have shown an even smaller proportion of COVID-19 cases with concurrent nasal congestion.^8^ Proposed mechanisms for COVID-19–related olfactory and taste dysfunction include an affinity for coronaviruses to infect olfactory nerves in an animal model and the broad expression of the receptor involved in the pathogenesis of SARS-CoV-2 on the epithelial cells of the tongue and oral cavity mucosa.^6,9,10^ With recent case series and case reports describing central and peripheral nervous system abnormalities, loss of taste and smell may represent only a subset of neurologic manifestations of COVID-19.^11,12^

This analysis is subject to limitations. All symptoms and medical histories were self-reported and limited by patient recall, health literacy (including younger children who may not have been able to adequately identify or describe symptoms), and availability of home thermometers. Symptoms prior to enrollment were collected retrospectively, and thus timelines for specific symptoms are not available. Because symptom ascertainment and SARS-CoV-2 testing were conducted at enrollment, any subsequent symptom development or detection of viral RNA that could affect PPVs for specific symptoms were not captured in this analysis. Prior reports have indicated that some symptoms, including the classic symptoms of fever and shortness of breath, may not present until later in the illness course.^13^ Due to the high prevalence of COVID-19 infection within this population, the PPV identified in this analysis may not be representative of all clinical encounters. Finally, the sample size was small and was not powered for detecting differences among subpopulations, and resulted in wide overlapping confidence intervals for the reported PPVs.

In this investigation, adding loss of taste and/or smell to the classic clinical criteria would have captured 95% of cases while only misidentifying two non-cases as cases, compared to 14 non-cases that would have been misidentified by the classic symptoms. CSTE recently added “new olfactory and taste disorder(s)” to its outpatient/telehealth clinical criteria for reporting.^4^ In the absence of confirmatory laboratory testing, criteria for a probable COVID-19 case now include loss of taste and/or smell in conjunction with other non-classic symptoms. Our early findings from a household transmission investigation suggest that adding loss of taste and/or smell to the singular CSTE clinical criteria, which currently include cough, shortness of breath, and difficulty breathing, may increase the efficiency of probable COVID-19 case identification.

Due to limited testing capacity, most states have prioritized testing of moderately to severely ill patients. However, as the availability of contact tracing and testing expands, testing and diagnoses will shift to also include outpatients with milder illness. Identifying these cases will assist in appropriate isolation recommendations and the prevention of additional spread within the community. Clinicians will benefit from further characterization of the full spectrum of illness in patients and may consider using loss of taste and/or smell in their testing strategies.

## Methods

All activities in this investigation were part of the public health response and were determined as not falling under human subjects research; data collection instruments were reviewed and approved by the Office of Management and Budget (OMB: 0920-1011). During March–April 2020, health department personnel in Milwaukee County, Wisconsin, assisted CDC in identifying a convenience sample of households through routine surveillance. Eligible households for the household study had a laboratory-confirmed COVID-19 index case ≤10 days from diagnosis who resided in the home at the time of enrollment and lived with at least one other household member. Informed consent was obtained from all participants, including index cases and their household members. Seven household members from two households chose not to participate. All participants completed questionnaires assessing basic demographic information, medical history, symptoms since the household index case’s disease onset, and other epidemiological information. A parent or legal guardian assisted in completing questionnaires for children <18 years. At enrollment, nasopharyngeal (NP) swabs were collected from all participants; to fulfill a secondary objective of this investigation, nasal self-collect (NSC) swabs were also collected from index cases and household members with classic symptoms of COVID-19 infection.

Swabs were tested by the City of Milwaukee Health Department Laboratory using the CDC SARS-CoV-2 real-time reverse transcription polymerase chain reaction (RT-PCR) assay.^14^ Any participant having a SARS-CoV-2–positive NP swab collected in the 10 days prior to enrollment (26 index cases and 2 household contacts) or a SARS-CoV-2–positive NP or NSC swab collected at enrollment was classified as a COVID-19 case.

Prevalence of symptoms among cases was compared descriptively. Bivariate analyses of demographic variables, medical history, and symptoms of reported loss of taste and/or smell were computed using two-tailed Pearson’s chi-squared test or Fisher’s exact test. To calculate the positive predictive value and 95% CIs of individual symptoms for COVID-19 among household contacts, we used any PCR-positive swab as the gold standard for diagnosis.. Statistical analyses were performed using SAS® Version 9.4, SAS Institute Inc., Cary, North Carolina.

## Disclaimer

The findings and conclusions in this report are those of the authors and do not necessarily represent the official position of the Centers for Disease Control and Prevention.

## Data Availability

Data presented in the current study may be available from the corresponding author on request.

## Acknowledgements

Households participating in the transmission study; Ann Christiansen, Kala Hardy, Christine Cordova, Kevin Rorabeck, and Kathleen Platt, North Shore Health Department; Jeanette Kowalik, Heather Paradis, Julie Katrichis, Catherine Bowman, Nancy Burns, Barbara Coyle, Elizabeth Durkes, Carol Johnsen, Jill LeStarge, Erica Luna-Vargas, Sholonda Morris, Mary Jo Gerlach, Jill Paradowski, Lindsey Page, Bill Rice, Michele Robinson, Virginia Thomas, Keara Jones, Chelsea Watry, and Richard Weidensee, City of Milwaukee Health Department; Jordan Hilleshiem, Beth Pfotenhauer, Manjeet Khubbar, Jennifer Lentz, Kristin Schieble, Noah Leigh, Joshua Weiner, Tenysha Guzman, Kathy Windham, and Julie Plevak, City of Milwaukee Health Department Laboratory SARS-CoV-2 testing team; Laura Conklin, Paige Bernau, and Emily Tianen, Wauwatosa Health Department; Wisconsin Department of Health Services COVID-19 response team; John Watson, Susan Gerber, Alicia Fry, and Aron Hall, CDC IMS COVID-19 Epidemiology Task Force; Daniel Owusu, Mary Pomeroy, Ashutosh Wadhwa, and Anna Yousaf, CDC COVID-19 field team.

## References

1. Guan, W., et al. Clinical Characteristics of Coronavirus Disease 2019 in China. N. Engl. J. Med. https://doi.org/10.1056/NEJMoa2002032 (2020).

2. Wang, D., et al. Clinical Characteristics of 138 Hospitalized Patients With 2019 Novel Coronavirus–Infected Pneumonia in Wuhan, China. JAMA 323, 1061–1069 (2020).

3. Centers for Disease Control and Prevention. Symptoms of Coronavirus. https://www.cdc.gov/coronavirus/2019-ncov/symptoms-testing/symptoms.html (2020).

4. Council of State and Territorial Epidemiologists. Technical Guidance Interim-20-ID-01: Standardized surveillance case definition and national notification for 2019 novel coronavirus disease (COVID-19). https://cdn.ymaws.com/www.cste.org/resource/resmgr/2020ps/interim-20-id-01_covid-19.pdf (2020).

5. Eliezer, M., et al. Sudden and Complete Olfactory Loss Function as a Possible Symptom of COVID-19. JAMA Otolaryngol. Head Neck Surg. https://doi.org/10.1001/jamaoto.2020.0832 (2020).

6. Giacomelli, A., et al. Self-reported Olfactory and Taste Disorders in Patients with Severe Acute Respiratory Coronavirus 2 Infection: A Cross-sectional Study. Clin. Infect. Dis. https://doi.org/10.1093/cid/ciaa330 (2020).

7. Westergaard, R. Urgent Update – Prioritization of COVID-19 Testing for Hospitalized Patients. https://www.dhs.wisconsin.gov/dph/memos/communicable-diseases/2020-09.pdf (2020).

8. Spinato, G., et al. Alterations in Smell or Taste in Mildly Symptomatic Outpatients with SARS-CoV-2 Infection. JAMA https://doi.org/10.1001/jama.2020.6771 (2020).

9. Netland, J., Meyerholz, D.K., Moore, S., Cassell, M., Perlman, S. Severe Acute Respiratory Syndrome Coronavirus Infection Causes Neuronal Death in the Absence of Encephalitis in Mice Transgenic for Human ACE2. J. Virol. 82, 7264–7275 (2008).

10. Xu, H., et al. High expression of ACE2 receptor of 2019-nCoV on the epithelial cells of oral mucosa. Int. J. Oral Sci. https://doi.org/10.1038/s41368-020-0074-x (2020).

11. Mao, L., et al. Neurologic Manifestations of Hospitalized Patients With Coronavirus Disease 2019 in Wuhan, China. JAMA Neurol. https://doi.org/10.1001/jamaneurol.2020.1127 (2020).

12. Schiedl, E., Canseco, D.D., Hadji-Naumov, A., Bereznai, B. Guillain-Barre syndrome during SARS-CoV-2 pandemic: a case report and review of recent literature. J. Peripher. Nerv. Syst. https://doi.org/10.1111/jns.12382 (2020).

13. Huang, C., et al. Clinical features of patients infected with 2019 novel coronavirus in Wuhan, China. Lancet 395, 497–506 (2020).

14. Centers for Disease Control and Prevention. Interim guidelines for collecting, handling, and testing clinical specimens from persons for coronavirus disease 2019 (COVID-19). https://www.cdc.gov/coronavirus/2019-nCoV/lab/guidelines-clinical-specimens.html (2020).

